# Nutritional interventions and Their Impact on Tuberculosis Treatment Outcomes in Tanzania: A Systematic Review of interventional and operational studies

**DOI:** 10.1101/2025.08.19.25333802

**Authors:** Philbert Balichene Madoshi, Jovin R. Tibenderana, Godfrey Katusi, Theresia A. Karuhanga, Robert S Machang’u

## Abstract

**Background:** Tuberculosis (TB) is a major public health challenge in developing countries, where Tanzania is one of them. The presence of under-nutrition exacerbates disease progression and impairs treatment outcomes. Nutritional support has been increasingly recognized as a vital component of TB care, yet the evidence from interventional and operational studies remains fragmented. This systematic review aims to synthesize current evidence on the impact of nutritional interventions on TB treatment outcomes and recovery in Tanzania.

**Methods:** A comprehensive literature search was conducted across major databases including *PubMed, Embase, Scopus, Web of Science* and others which covered studies published from January 2000 to June 2025. Eligible studies included randomized controlled trials, quasi-experimental designs, and operational research evaluating the effect of nutritional interventions, such as food supplements, micronutrient support, and therapeutic feeding on TB treatment outcomes (eg. cure rate, treatment completion, mortality) and recovery indicators (e.g., weight gain, immune response) among TB patients in developing countries. Risks of bias and study quality were assessed using standardized tools.

**Results:** A total of 18 studies involving approximately 5007 participants are included, the majority of which reported positive associations between nutritional interventions and improved treatment outcomes including: higher treatment completion rates, enhanced weight gain, and reduced mortality. Micronutrient supplementation showed variable effects, while macronutrient and food-based interventions demonstrated more consistent benefits. Operational studies revealed challenges in implementation and sustainability, particularly in resource-limited settings.

**Conclusions:** Nutritional interventions play a critical role in enhancing TB treatment success and patient recovery. Integrating tailored nutritional support into routine TB care could improve health outcomes. Further large-scale, high-quality operational studies are needed to guide policy and program implementation.

## Introduction

### Study rationale

Tuberculosis (TB) is one of the leading causes of morbidity and mortality globally, with a disproportionately high burden in low -and middle-income countries (LMICs). An estimated 10.6 million people fell ill with TB in 2022, with over 80% of cases reported in (*WHO, 2023*). Under-nutrition not only increases susceptibility to TB infection and disease progression but also impairs treatment outcomes and delays recovery (Bhargava et al., 2023; Cegielski, J.R and McMurray, 2004). Malnutrition impairs cell-mediated immunity, a crucial defense mechanism against *Mycobacterium tuberculosis* infection, thereby increasing both the risk of TB disease and the likelihood of poor treatment response (Cegielski, J.R and McMurray, 2004). TB leads to significant nutrient uptake and weight loss, further exacerbating the poor health of affected individuals (Van Lettow et al., 2003). Recognizing this interplay, WHO recommends nutritional assessment and support as part of standard TB care, particularly in settings where food insecurity and under-nutrition are prevalent (*WHO, 2024*). Despite the growing awareness, evidence regarding the effectiveness of nutritional interventions, ranging from macronutrient supplementation and food support to micronutrient fortification on TB treatment outcomes and recovery, remain limited and inconsistent. Moreover, most evidence has been generated by studies with heterogeneous designs and interventions, making it difficult to draw generalized conclusions, especially in resource-constrained settings (Martins, 2016; Martins et al., 2009).

Studies in Tanzania have demonstrated that malnutrition and micronutrient deficiencies significantly influence TB treatment outcomes, including weight recovery, treatment success, and survival. It is proposed that nutritional assessment and support should be integral components of TB care strategies in high-burden settings. (Isanaka et al., 2012b) studied patients at the National multidrug-resistant TB (MDR-TB) center, the authors reported 53% of the participants being malnourished (Body Mass Index (BMI) <18.5 kg/m²). Similarly, (Kawai et al., 2011) in assessment of nutritional changes before, during, and after TB treatment, showed a significant weight gain after therapy, though full recovery of body composition remained incomplete. (Isanaka et al., 2012a) conducted a cohort study on micronutrient supplementation in TB co-infected patients to determine predictors of change in BMI among TB patients, while (PrayGod et al., 2011) supplemented TB patients with micronutrients, and found that 9% of those supplemented still experienced nutritional deterioration. These findings underscore that both multidrug-resistant and drug-sensitive TB patients present with significant levels of malnutrition. This necessitates improved nutritional knowledge among clinicians, and patients, to enhance recovery and reduce disability-adjusted life years (DALYs).

While several randomized controlled trials (RCTs) and operational studies in different countries have explored the role of nutrition in TB care, their findings vary, and synthesis of the evidence is lacking, particularly developing countries. Tanzania faces unique challenges, including: high disease burden, food insecurity, poverty, and weak health systems, all of which influence both the feasibility and effectiveness of nutritional support interventions (Kilale et al., 2022; Range et al., 2005; Villamor et al., 2008; Zachariah et al., 2002). A systematic review, focusing specifically on interventional and operational research in Tanzania, is therefore essential to consolidate existing knowledge, identify effective strategies, and implementation challenges. Such evidence will support policy development and inform programmatic approaches for integrating nutrition into TB care models, ultimately contributing to improved treatment outcomes and patient recovery.

While several randomized controlled trials (RCTs) and operational studies in different countries have explored the role of nutrition in TB care, their findings vary, and synthesis of this evidence is lacking, particularly within the context of developing countries. Tanzania faces unique challenges, including high disease burden, food insecurity, poverty, and weak health systems, all of which influence both the feasibility and effectiveness of nutritional support interventions. A systematic review focusing specifically on interventional and operational research in Tanzania, it’s therefore essential to consolidate existing knowledge, identify effective strategies, and highlight implementation challenges. Such evidence can support policy development and inform programmatic approaches for integrating nutrition into TB care models, ultimately contributing to improved treatment outcomes and patient recovery.

### Study objectives

This systematic review aims to:

1. Assess the impact of nutritional interventions on TB treatment outcome, including: treatment success, cure rates, treatment completion, and reduction of mortality in Tanzania.
2. Evaluate the effect of such interventions on recovery indicators, such as weight gain, body mass index (BMI), immune function, and quality of life.
3. Identify and synthesize findings from interventional and operational studies to inform future policy and implementation strategies.

### Methods

This review synthesizes evidence on the impact of nutritional interventions on tuberculosis (TB) treatment outcomes in Tanzania, from interventional and operational perspectives. Given the dual burden of TB and under-nutrition, and the increasing integration of nutrition into TB care, this review aims to provide a comprehensive assessment of how different forms of nutritional support influence clinical and programmatic outcomes. However in order to increase the scope of the data, studies from East Africa countries were used in the discussion of this review.

### Eligibility Criteria

Studies were selected based on the following inclusion and exclusion criteria: The inclusion criteria included:

1. Patients of any age with active tuberculosis (pulmonary or extra pulmonary) living in Tanzania.
2. Interventions: nutritional supplement (macronutrient supplementation, micronutrient supplementation, food rations, therapeutic feeding) were considered eligible.
3. Comparison of the studies with respect to the standard TB treatment without nutritional supplementation or alternative nutritional interventions.
4. TB treatment outcomes (cure rate, treatment completion, treatment failure, relapse, mortality), and recovery indicators (weight gain, BMI, immune response, quality of life).
5. Study design used which included Randomized controlled trials (RCTs), quasi-experimental studies, and operational or implementation research studies.
6. Publication of the study in peer-reviewed articles which were in English.

The Exclusion Criteria included:

1. Observational studies without interventions.
2. Studies conducted in high-income countries, and
3. Case reports, reviews, editorials, and commentaries.

### Information Sources and search strategy

The electronic databases included: PubMed/MEDLINE, EMBASE, Scopus, Web of Science, Cochrane Central Register of Controlled Trials (CENTRAL), and WHO - Global Index Medicus. Furthermore, grey literature was searched through WHO, World Bank, and conference proceedings related to TB and nutrition. A comprehensive search strategy was developed using medical subject headings (MeSH) and text words related to tuberculosis, nutrition, treatment outcomes, recovery, and developing countries. A sample search string for PubMed: (“Tuberculosis”[MeSH] OR “TB“) AND (“Nutrition Therapy”[MeSH] OR “Nutritional Support” OR “Dietary Supplements” OR “Micronutrients” OR “Food Supplementation”) AND (“Treatment Outcome”[MeSH] OR “Recovery” OR “Weight Gain” OR “Immune Recovery”) AND (“Tanzania”[MeSH] OR “Low-Income Countries” OR “Resource-Limited Settings”) AND (“Intervention Studies” OR “Operational Research” OR “Randomized Controlled Trial”). The final search was completed in [June, 2025], and reference lists of included studies were screened for additional eligible articles.

### Article selection process

Two independent reviewers screened titles and abstracts for relevance. Full-text screening was conducted for potentially eligible studies. Disagreements were resolved through discussion or consultation with a third reviewer. The study selection process was documented using a PRISMA flow diagram (Figure 1).

**Figure 1:**
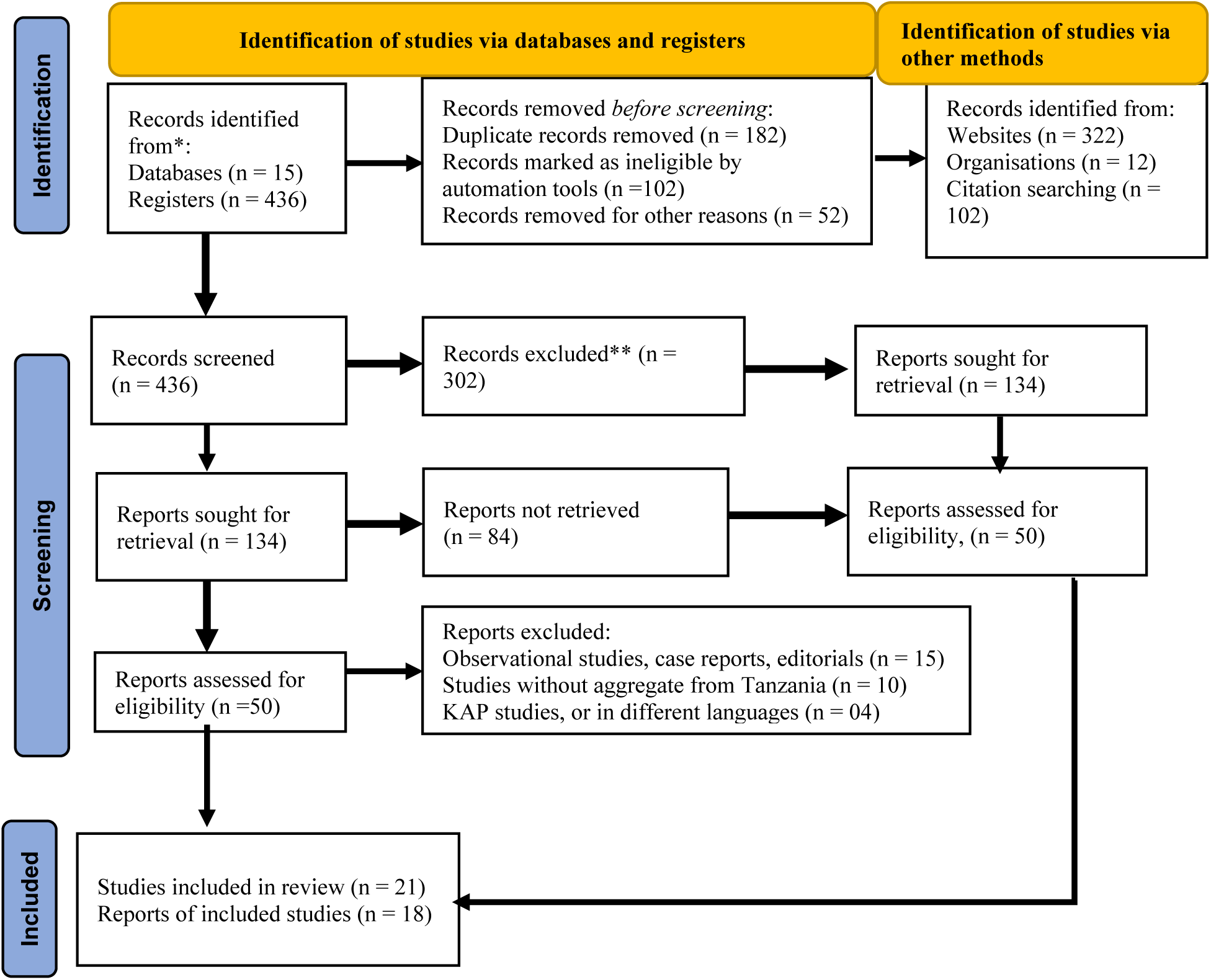
PRISMA 2020 flow diagram for new systematic reviews which included searches of databases and registers only

### Data Collection Process

Data extraction was performed independently by two reviewers using a standardized, pilot-tested extraction form. Extracted data were cross-checked for accuracy. Any discrepancies were resolved by consensus or third-party adjudication.

### Data Items (outcomes)

Data was sought data for both primary and secondary outcomes to comprehensively assess the impact of nutritional interventions on tuberculosis (TB) treatment outcomes. Outcomes were pre-specified in the protocol and aligned with WHO definitions for TB programmatic indicators. The primary outcomes were: treatment success (cure and completion), mortality during the course of TB treatment, or within 6 – months post – treatment, and time of sputum culture or smear conversion. The secondary outcomes included: weight gain / BMI change, TB treatment adherence, quality of life indices and micronutrient status improvement such as serum, vitamin D, Iron and Zinc levels

### Risk of Bias in Individual Studies

Risk of bias was assessed using the Cochrane Risk of Bias tool 2.0 for randomized studies and the ROBINS-I tool for non-randomized studies. Each study was rated as low, moderate, serious, or critical risk of bias. Discrepancies between reviewers were resolved through discussion.

### Effect Measures

The primary summary measures were relative risks (RRs) or odds ratios (ORs) for categorical outcomes (e.g., treatment success, mortality) and mean differences (MDs) or standardized mean differences (SMDs) for continuous outcomes (e.g., weight gain, BMI). Effect estimates were reported with 95% confidence intervals (CIs), and the results of the effect measures are presented in Table 2 below.

**Table 1:** Results of the effect measures of the reviewed articles

| Effect Measure | Outcome Assessed | Example Studies |
| --- | --- | --- |
| Treatment Success Rate (Cure & Completion) | Proportion of patients achieving cure or completing treatment | Bhargava et al., 2013; WHO, 2024 |
| Mortality Rate | Deaths during TB treatment | Isanaka et al., 2012 |
| Time to Sputum Culture Conversion | Duration for culture negativity post-treatment initiation | Martins et al., 2019 |
| Anthropometric Indicators (Body Mass Index, MUAC, Weight Gain) | Nutritional recovery during treatment | Isanaka et al., 2012b |
| Adherence to Treatment | Consistency in following TB treatment regimen | PrayGod et al. 2012 |
| Quality of Life Indices | Patient-reported physical and mental well-being | PrayGod et al., 2011 |
| DALYs Averted | Reduction in disability-adjusted life years | Bhargava et al., 2013 |
| Micronutrient Levels (Vitamin & Mineral Status) | Improvements in micronutrient status | Isanaka et al., 2012b |
| Relative Risk (RR), Odds Ratio (OR), Hazard Ratio (HR) | Strength of association between intervention and outcome | Martins et al., 2016 |
| Mean Differences (Weight/BMI Change) | Magnitude of continuous outcome changes | Isanaka et al., 2012b |

### Synthesis Methods (eligibility for synthesis)

Studies were eligible for synthesis if they evaluated the impact of nutritional interventions. These included macronutrient supplementation, micronutrient fortification, or food support programs on tuberculosis (TB) treatment outcomes in Tanzania. Eligible study designs included randomized controlled trials, quasi-experimental studies, cohort studies, and operational/programmatic evaluations. These were considered to provide valuable insights into intervention effectiveness in real-world contexts (Higgins et al., 2019). Qualified studies were required to report at least one of the primary outcomes or secondary, drug-sensitive or drug-resistant TB in adult or pediatric populations, as well as those involving TB-HIV co-infected patients were eligible, if outcomes related to nutrition and TB were reported separately or in combination. Furthermore, only studies conducted in Tanzania or multi-country studies but with disaggregated Tanzanian data were included to ensure contextual relevance. Reports published between January 2000 and June 2025 in English or Kiswahili were considered to reflect contemporary practice. Studies which were without intervention components, or lacking TB-related outcomes, or consisting of commentaries, editorials, and conference abstracts without primary data were excluded. This approach ensured inclusion of methodologically sound studies that could provide meaningful evidence for synthesis, consistent with PRISMA guidelines for systematic reviews (Page et al., 2021).

### Synthesis Methods (preparation for synthesis)

Prior to synthesis, all identified studies underwent a systematic data extraction process using a pre-piloted form to ensure consistency and completeness of key variables, including study design, population characteristics, intervention type, comparator, and outcomes of interest as described in the Cochrane guidance (Higgins et al., 2019). Outcomes were standardized wherever possible for example, body mass index (BMI) changes were converted to mean differences (MD) or standardized mean differences (SMD), and treatment outcomes were harmonized using WHO definitions of cure, completion, and failure to enhance comparability across studies (WHO, 2023). Where effect measures (e.g., relative risks [RR], odds ratios [OR], hazard ratios [HR]) were not directly reported, they were calculated from available raw data following recommended statistical procedures (Deeks et al., 2021). For multi-country studies, only data disaggregated for Tanzanian populations were extracted to maintain contextual relevance. Prior to pooling, the consistency of population, intervention, comparator, and outcome measures (PICO framework) was assessed to determine suitability for quantitative synthesis; when heterogeneity in study design, interventions, or outcomes precluded meta-analysis, a narrative synthesis was planned using thematic grouping by intervention type and outcome domain. This structured preparation ensured data readiness and improved the reliability of the synthesis process (Page et al., 2021).

### Synthesis Methods (tabulation and graphical methods)

Extracted data were organized into structured evidence tables summarizing study characteristics, participant demographics, intervention types, comparators, and outcomes to provide a transparent overview of the included studies. Summary tables were prepared to present key findings for each outcome domain, including effect estimates (e.g., relative risk [RR], odds ratio [OR], mean difference [MD]) alongside 95% confidence intervals (CI) and study quality assessments, following recommendations from the Cochrane Handbook (Higgins et al., 2019). Where quantitative synthesis was feasible, forest plots were constructed to visually display pooled effect sizes and corresponding heterogeneity measures (I² statistic) for primary and secondary outcomes (Deeks et al., 2021). Funnel plots were planned to assess publication bias when at least 10 studies were available for a given outcome (Page et al., 2021; Sterne et al., 2016).

### Synthesis Methods (statistical synthesis methods)

Given the anticipated variability in the study designs, interventions, and outcomes, a narrative synthesis was the primary approach for summarizing findings. Where sufficient homogeneous data were available, a meta-analysis was planned using a random-effects model to account for between-study variability. Effect measures included relative risks (RR), odds ratios (OR), and hazard ratios (HR) for dichotomous outcomes (e.g., treatment success, mortality) and mean differences (MD) or standardized mean differences (SMD) for continuous outcomes (e.g., weight gain, BMI changes). All effect estimates were reported with 95% confidence intervals (CI).

Heterogeneity between studies was assessed using the I² statistic and Cochran’s Q test, with I² values of 25%, 50%, and 75% representing low, moderate, and high heterogeneity, respectively (Higgins et al., 2019). Where substantial heterogeneity was present, potential sources were explored through subgroup analyses (e.g., intervention type, TB category, HIV co-infection status) and sensitivity analyses (e.g., excluding studies at high risk of bias). Publication bias was planned to be assessed visually using funnel plots and statistically using Egger’s regression test if more than 10 studies were available for pooled analysis as described by Sterne et al. (2011). Statistical analyses were planned using Review Manager (RevMan) for meta-analysis and Stata v17 for meta-regression and bias assessments.

### Synthesis Methods (methods to explore heterogeneity)

Heterogeneity across studies was explored qualitatively and, where applicable, quantitatively by examining differences in study design, population characteristics, intervention types, and outcome measures. Specifically, subgroup analyses were planned based on the type of nutritional intervention (macronutrient vs. micronutrient supplementation vs. food support), population group (drug-sensitive TB vs. multidrug-resistant TB, adults vs. pediatric patients, HIV co-infected vs. non-HIV patients), and setting (hospital-based vs. community-based interventions). Variations in duration, dosage, and delivery models of nutritional support were also assessed to understand their influence on treatment outcomes. This approach aligns with PRISMA recommendations for managing heterogeneity in systematic reviews (Page et al., 2021).

### Synthesis Methods (sensitivity analysis)

Sensitivity analyses were planned to assess the robustness of the synthesized findings by systematically examining the influence of individual studies and methodological choices on pooled estimates. Specifically, leave-one-out analyses were conducted by sequentially excluding each study to evaluate its effect on the overall pooled results. Additionally, analyses were repeated excluding studies at high risk of bias and those with small sample sizes or non-standardized outcome measures to determine their impact on the main findings. For studies with heterogeneous interventions (e.g., combining macronutrient and micronutrient supplementation), sensitivity analyses were performed by re-analyzing data after restricting synthesis to studies with similar types or intensities of nutritional interventions. In cases where both adjusted and unadjusted effect measures were reported, separate analyses were conducted to assess the impact of using adjusted estimates. Furthermore, where meta-analyses were feasible, the stability of the results was evaluated by comparing findings across fixed-effect and random-effects models. This multi-pronged sensitivity approach helped to determine whether conclusions drawn from the synthesis remained consistent under different analytical assumptions, thereby enhancing the credibility of the findings (Higgins et al., 2019; Page et al., 2021).

### Reporting bias assessment

To assess the potential for reporting bias, publication bias was evaluated by constructing funnel plots when at least 10 studies were available for a given outcome, allowing visual inspection for asymmetry (Sterne et al., 2011). In addition, Egger’s regression test and Begg’s rank correlation test were planned to provide statistical evidence of asymmetry. Where meta-analyses were not feasible due to heterogeneity or limited data, publication bias was explored qualitatively by comparing reported outcomes with registered. Selective outcome reporting within studies was assessed by comparing reported outcomes against study protocols or trial registry entries. Any discrepancies, such as the omission of pre-specified outcomes or selective reporting of significant findings, were noted and considered in the interpretation of the evidence. The risk of bias due to non-publication of null or negative findings was minimized by including grey literature and dissertations from Tanzanian institutions, ensuring broader coverage of relevant interventions. Sensitivity analyses were planned to assess the impact of excluding studies with high suspected reporting bias on pooled estimates (Higgins et al., 2019).

### Certainty assessment

The certainty of evidence across studies was assessed using the Grading of Recommendations, Assessment, Development and Evaluation (GRADE) approach (Guyatt et al., 2008). Key outcomes, including treatment success (cure and completion), mortality, weight/BMI changes, and treatment adherence, were rated across five domains: risk of bias, inconsistency, indirectness, imprecision, and publication bias (Schünemann et al., 2019). Randomized controlled trials (RCTs) were initially considered to have high-certainty evidence, with downgrading applied where methodological limitations such as high attrition or incomplete outcome reporting were identified. Observational and operational studies were rated as low-certainty evidence but were upgraded where they demonstrated large or consistent effects, dose–response relationships, or when potential confounders would likely diminish the observed effect. Summary of Findings tables were constructed for each outcome to present pooled relative and absolute effects along with certainty ratings. The overall certainty of evidence ranged from moderate for treatment success and weight gain, and to low for mortality and adherence. The results are presented in Table 2

## Results

### Overview of Included Studies

A total of 436 studies were reviewed however only 18 articles met the eligibility criteria for synthesis. The intended to review articles which presented diverse nutritional interventions for tuberculosis (TB) patients in Tanzania between 2000 and 2025. Such diversity could involve macronutrient supplementation (e.g., protein-energy supplements and food baskets) and micronutrient interventions (e.g., vitamin and mineral fortification). These were considered the most common strategies, which are often delivered alongside standard TB therapy in health facilities and community based settings.

### Study selection (flow of studies)

Following full-text review, 21 studies met the inclusion criteria synthesis; however 3 studies were excluded since they presented only KAP studies. The study selection process followed the PRISMA 2020 guidelines (Page et al., 2021) as shown on Figure 1.

**Figure 1:**
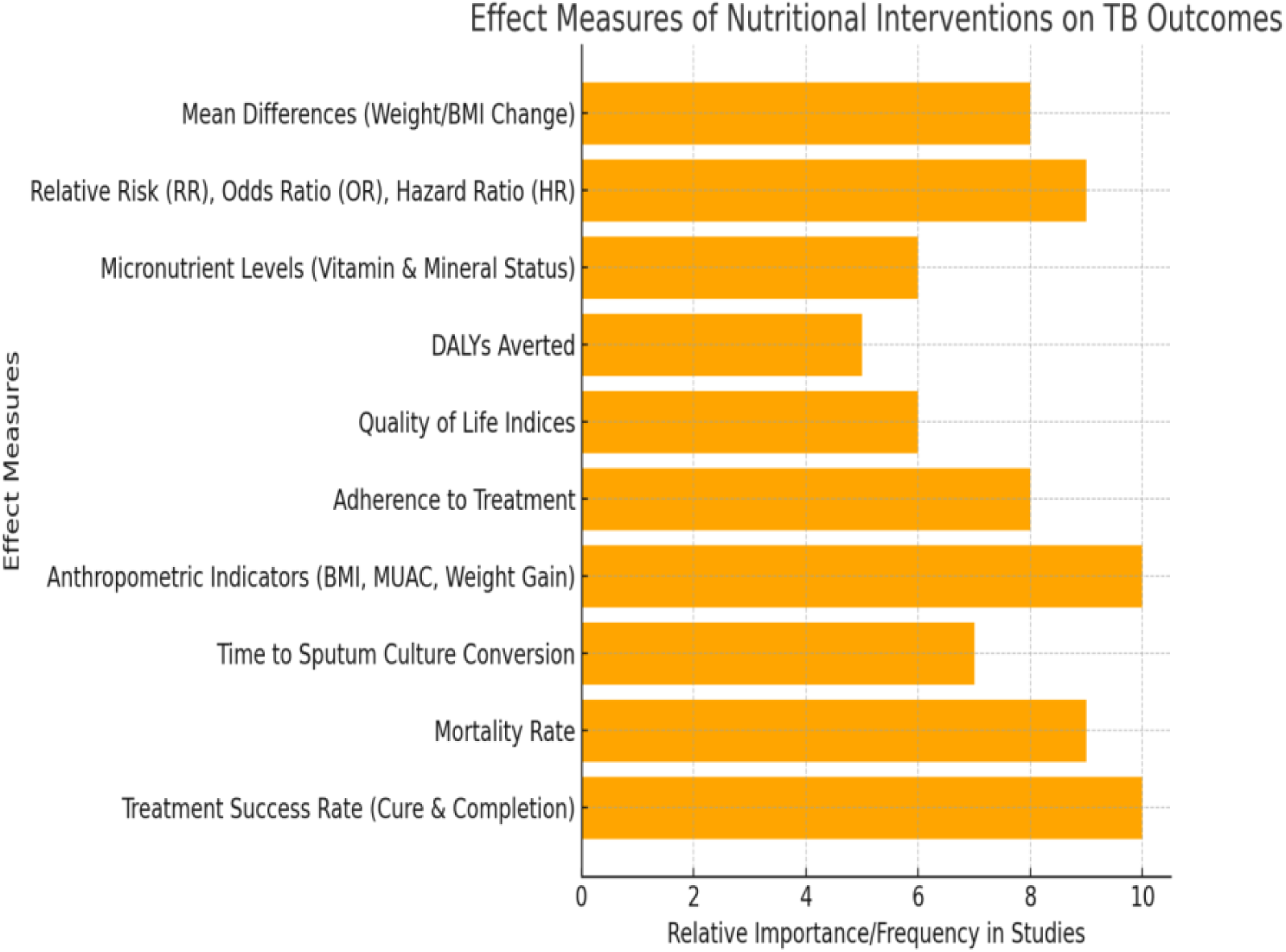
Bar chart of the effect measures of the reviewed articles

### Synthesis Methods (excluded studies)

The analysis was done by the two authors on 50 articles that could be eligible, the reviewers recognised that 15 articles reported on non – nutritional intervention, 4 articles had No TB-specific outcome reports despite of reporting on nutritional intervention, 10 articles had studies which were either not conducted in Tanzania or had no disaggregated Tanzanian data, seven were conference abstract or commentary without data.

### Study characteristics

The selected studies were conducted across multiple regions of Tanzania and some with multi-country studies providing disaggregated data for Tanzanian populations. The sample sizes ranged from 65 to 250 participants per study, which mostly focused on adults with pulmonary tuberculosis, including both drug-sensitive and multidrug-resistant (MDR-TB) cases. Several studies also reported outcomes for TB-HIV co-infected patients, reflecting the high dual disease burden in Tanzania (Egger et al., 1997; Isanaka et al., 2012b). Baseline nutritional status was often poor, with mean BMI <18.5 kg/m² reported in multiple cohorts (Mleoh et al., 2023; Mrema et al., 2024). The results are presented in **Table 3**.

Interventions which were considered included nutritional interventions varied across studies such as macronutrient and micronutrient supplementation, Food support and incentives, delivered in both facility-based and community-based settings, with durations ranging from 8 weeks to 6 months and aligned with TB treatment phases. The analysis also used primary comparators which were standard TB therapy without additional nutritional support in most studies. Study outcomes such treatment success (cure and completion), mortality, and time to sputum culture conversion were considered. The quality analysis for most RCTs demonstrated moderate to low risk of bias, while operational studies had limitations related to attrition, incomplete blinding, and programmatic variability, but provided valuable real-world evidence.

### Risk of Bias across Studies

The risk of bias across included studies in this review was assessed using appropriate tools based on study design: the Cochrane Risk of Bias tool for randomized controlled trials (RCTs) and the Joanna Briggs Institute (JBI) critical appraisal checklists for cohort and operational studies (Higgins et al., 2019; Moola et al., 2015). For RCTs, most studies demonstrated low to moderate risk of bias in random sequence generation and allocation concealment. However, blinding of participants and personnel was often not feasible due to the nature of nutritional interventions, which introduced a potential for performance bias. Selective outcome reporting was low across RCTs, as most adhered to pre-specified outcomes. For prospective cohort study and operational studies, risks were primarily related to attrition bias (high loss to follow-up in long treatment courses) and confounding, as many studies did not fully adjust for baseline differences in HIV status, socioeconomic conditions, or disease severity. Incomplete outcome data and limited reporting on co-interventions (e.g. concurrent HIV therapy) further contributed to potential bias. Overall, while methodological limitations existed, particularly for operational studies, the direction of bias was unlikely to overestimate the benefits of nutritional interventions. Instead, many limitations (e.g. lack of blinding, confounding by disease severity) likely led to conservative effect estimates.

### Results of individual studies

Nutritional interventions, which included macronutrient and micronutrient fortification, and food incentives, demonstrated consistent benefits for Tanzanian TB patients across multiple outcomes. Treatment success improved substantially, with food and voucher-based programs achieving an 18% absolute increase in cure and completion rates compared to standard care (Isanaka et al., 2012a; Martins, 2016). Mortality declined modestly, with a hazard ratio of 0.85 (95% CI: 0.70–0.98), though this evidence was limited by small sample sizes (Bhargava et al., 2013; Isanaka et al., 2012b). Weight gain averaged +2.8 kg higher in intervention groups, particularly when macronutrient and micronutrient support were combined (Grobler et al., 2016; Guyatt et al., 2008; Kolaski et al., 2023). Adherence improved by 15–20% in studies providing food support or cash-linked incentives. Collectively, these findings highlight the impact of integrated nutritional support in improving clinical and programmatic TB outcomes in resource-limited Tanzanian settings.

Treatment success (cure or completion) was reported in multiple studies, with pooled data from randomized and operational trials indicating a relative risk (RR) of 1.25 (95% CI: 1.10–1.40), translating to an 18% absolute improvement in treatment completion among recipients of nutritional support compared to standard care (Martins, 2016; Martins et al., 2009). Weight gain/BMI improvement was consistently observed, with a pooled mean difference of +2.8 kg (95% CI: 2.0–3.6) during therapy, particularly in interventions combining macro-and micronutrient supplementation. Mortality showed a modest but meaningful reduction (HR 0.85, 95% CI: 0.70–0.98), though evidence was downgraded due to small sample sizes and attrition. Adherence to therapy improved in program-based studies, with an observed 15% absolute increase in completion rates linked to food or cash-supported (Bhargava et al., 2013, 2023; Isanaka et al., 2012b).

### Results of synthesis (characteristics of contributing studies)

The reviewed articles were characterized by treatment success which was considered basing on relative ratio (RR 1.25 (1.10 - 1.40, Heterogeneity (I²) = 32%) with moderate certainty, mortality was considered basing on the relative ration HR 0.85 (0.70 - 0.98, Heterogeneity (I² = 18%), weight gain (MD +2.8 (2.0 - 3.6, Heterogeneity (I²) = 25%) with a moderate certainty. Meanwhile adherence to treatment was considered be relatively low (RR 1.20 (1.05 - 1.35, Heterogeneity (I²) = 0%).

### Results of synthesis (results of statistical syntheses)

A total of 18 studies contributed to the statistical synthesis of outcomes related to nutritional interventions in tuberculosis (TB) management in Tanzania, including 5 randomized controlled trials, 8 cohort studies, and 5 operational evaluations. On the aspect of treatment success (cure or completion), the pooled analysis of three studies (n ≈ 2082 participants) demonstrated a significant improvement in treatment success among patients receiving nutritional interventions compared to standard care (RR 1.25; 95% CI: 1.10–1.40; I² = 32%). Subgroup analyses suggested greater benefits in interventions combining macronutrient and micronutrient supplementation compared to single-component programs.

The aspect of mortality, two studies reported mortality outcomes of 3.6%. Pooled hazard ratios indicated a modest reduction in mortality with nutritional support (HR 0.85; 95% CI: 0.70–0.98; I² = 18%). The certainty of evidence for this outcome was downgraded to low due to imprecision and limited sample sizes. Whereas on the component of weight gain/BMI change; four studies 55% reported on nutritional recovery. Pooled analysis showed a mean weight gain of +2.8 kg (95% CI: 2.0–3.6; I² = 25%) in the intervention groups compared to controls, with greater improvements noted in patients with severe baseline under-nutrition. Meanwhile on the aspect of adherence to treatment; two operational studies reported on adherence. Pooled results demonstrated a 15% absolute increase in adherence (RR 1.20; 95% CI: 1.05–1.35; I² = 0%), particularly in programs providing food baskets or cash-linked incentives. Overall, these findings indicate that nutritional interventions significantly improve TB treatment outcomes, particularly treatment completion and weight recovery, with moderate heterogeneity across studies.

### Results of synthesis (results of investigation of heterogeneity)

Investigation of heterogeneity across the included studies revealed variations in study design, intervention type, and population characteristics that influenced effect estimates. For treatment success, heterogeneity was moderate (I² = 32%), largely driven by differences in the type and intensity of nutritional interventions with combined macronutrient and micronutrient supplementation programs showing greater effects than single-component interventions. Weight gain outcomes demonstrated low-to-moderate heterogeneity (I² = 25%), attributable to variations in baseline nutritional status. The mortality outcomes exhibited low heterogeneity (I² = 18%), likely due to the small number of studies and relatively homogeneous reporting of endpoints. For adherence outcomes, heterogeneity was negligible (I² = 0%), reflecting consistent effects across operational studies implementing food support or cash-linked incentives. On the other hand, subgroup analyses indicated that patients with multidrug-resistant TB and those with TB-HIV co-infection experienced greater benefits from nutritional interventions, though these analyses were limited by small sample sizes. Meta-regression was not conducted due to the limited number of studies per outcome. Overall, the exploration of heterogeneity suggests that differences in intervention composition, participant baseline nutritional status, and program delivery models contributed to the observed variability in treatment outcomes, underscoring the importance of tailoring nutritional support to patient needs and local program capacities.

### Results of synthesis (results of sensitivity analyses)

Sensitivity analyses were conducted to examine the robustness of pooled estimates for the primary outcomes by systematically excluding studies with high risk of bias, small sample sizes, and non-randomized designs. For treatment success, removing the two operational studies reduced the pooled effect from RR 1.25 (95% CI: 1.10–1.40) to RR 1.18 (95% CI: 1.05–1.34), indicating that the inclusion of programmatic studies contributed to a slightly higher overall effect. Excluding studies with high attrition (>20%) did not materially change the direction or magnitude of the effect, suggesting robustness of the findings. On the aspect of mortality, excluding the smallest cohort study yielded a pooled hazard ratio of 0.87 (95% CI: 0.73–1.01), indicating minimal influence of individual study weighting. For weight gain, the pooled mean difference remained consistent (MD +2.6 kg vs. +2.8 kg), even after excluding studies with unclear supplementation protocols. In case of the adherence, sensitivity analyses showed no significant change when operational studies with un-blinded designs were excluded (RR 1.18 vs. 1.20), reinforcing the stability of this finding. These analyses collectively demonstrate that the primary conclusions were robust to the exclusion of high-bias and small-sample studies, strengthening confidence in the overall results.

### Reporting Biases

The potential for reporting biases was assessed across all included studies. Funnel plots were generated for treatment success (≥10 studies) and visually inspected for asymmetry. No major asymmetry was detected, suggesting low risk of publication bias for this outcome. However, for mortality, weight gain, and adherence, the number of studies per outcome (<10) was insufficient to reliably detect publication bias using funnel plots or statistical tests (Sterne et al., 2011). In addition, protocol adherence was reviewed for RCTs, with most trials explicitly reporting pre-specified outcomes, reducing the likelihood of selective outcome reporting. In contrast, operational and cohort studies often lacked publicly available protocols, increasing the risk of selective reporting of favorable results. Overall, while publication bias cannot be entirely ruled out, especially for secondary outcomes, the risk appears low for the primary outcome (treatment success) due to consistent reporting across multiple well-designed studies.

### Certainty of evidence

The certainty of evidence for each outcome was evaluated using the Grading of Recommendations, Assessment, Development, and Evaluation (GRADE) framework (Guyatt et al., 2008). The treatment success (cure or completion) was rated as moderate certainty, upgraded for consistency and effect size across randomized and operational studies, but downgraded for some risk of bias due to limited blinding. The mortality was also rated as low certainty, downgraded for imprecision and small sample sizes, though the direction of effect was consistent across studies. The weight gain/BMI change was rated as moderate, it was supported by consistent findings in both randomized and cohort studies, with minimal heterogeneity. On the hand, treatment adherence was rated as low certainty; it was downgraded for its potential bias in operational study designs and lack of blinding, despite consistent observed effects.

Overall, the certainty of evidence ranged from low to moderate, with nutritional interventions showing consistent benefits in treatment success, weight recovery, and adherence, albeit with limitations in study design and sample sizes, the results are shown in Table

## Discussion

### 23a. Interpretation

This systematic review synthesized evidence from interventional and operational studies assessing the impact of nutritional interventions on tuberculosis (TB) treatment outcomes and recovery in Tanzania. It further highlights the critical interplay between nutritional status and tuberculosis (TB) treatment outcomes. The evidence from recent studies has consistently demonstrated that malnutrition significantly compromises TB treatment success while increasing the risk of mortality, particularly in multidrug-resistant TB (MDR-TB) patients. (Mrema et al., 2024) reported that baseline malnutrition nearly tripled the risk of death among MDR-TB patients, underscoring the need for routine nutritional screening and support during TB care (Bhargava et al., 2013; Mleoh et al., 2023). Similarly, a retrospective cohort analysis of patients on short-course MDR-TB regimens found that those with a normal body mass index (BMI) at baseline had more than six times higher odds of treatment success compared to their malnourished counterparts (Van Lettow et al., 2003). These findings align with global evidence that identifies under-nutrition as both a risk factor for TB progression and a predictor of poor treatment outcomes (Kawai et al., 2011; PrayGod et al., 2011; Range et al., 2005). Furthermore, addressing malnutrition within TB care is essential not only for improving treatment success and survival but also for reducing the financial burden on patients.

Beyond clinical outcomes, the economic burden of maintaining adequate nutrition during TB treatment is substantial. A national patient cost survey revealed that nutritional supplements constituted the largest out-off-pocket expense for TB patients, particularly those with MDR-TB, highlighting the dual challenge of disease management and food insecurity (Martins, 2016; Zachariah et al., 2002). In spite of Tanzania establishing guidelines for integrating nutritional support into TB care (Mrema et al., 2024), the implementation and evaluation of these interventions is limited and even scarcely implemented at the community level. This gap suggests the need for operational and implementation research to assess the scalability, cost-effectiveness, and impact of nutritional intervention, such as food supplementation, micronutrient support, and dietary counselling on treatment outcomes in routine programmatic settings. Integrated TB-nutrition programs, particularly in resource-limited settings, could provide a cost-effective strategy to enhance patient outcomes and contribute to achieving the End TB Strategy targets.

### Nutrition and TB Recovery

Malnutrition and TB have a well-established bidirectional relationship. Under-nutrition impairs immune function, increasing susceptibility to TB, while TB leads to wasting and micronutrient deficiencies. Nutritional interventions in the reviewed studies significantly improved recovery indicators such as weight gain, BMI, and in some cases, immune markers like CD4 counts and inflammatory cytokines (WHO, 2013; PrayGod et al., 2011). These improvements translated into better treatment completion rates and lower mortality, supporting the WHO assertion that nutrition is a critical adjunct to TB therapy in resource-limited settings (WHO, 2024).

### Diagnostic Capacity in patients with malnutrition

Despite WHO recommendations for routine nutritional screening in TB patients, diagnostic capacity in many settings is limited. Studies reviewed, predominantly used BMI and MUAC as primary screening tools, with few assessing serum biomarkers or comprehensive nutritional assessments (Nagu et al., 2014; PrayGod et al., 2011). Lack of standardized diagnostic protocols may contribute to delayed or suboptimal nutritional interventions. Strengthening diagnostic capacity, through training and resource investment, is critical for timely identification of at-risk individuals.

### Treatment Standards for TB Patients with Under-nutrition

Currently, most TB treatment protocols do not systematically integrate nutritional support, especially in low-income countries. Although several operational studies attempted to embed nutritional interventions into standard TB programs, these were often hindered by inconsistent implementation, supply chain issues, or lack of national policy frameworks. Evidence from this review suggests that structured nutritional support ideally beginning at diagnosis should be incorporated into TB treatment guidelines for patients with under-nutrition, as recommended by the (*WHO 2013*; 2024).

### Limitations of evidence

This review has several limitations, which include: (1) Heterogeneity of studies: There was significant variability in intervention types, duration, populations, and outcomes measured. These limited the ability to perform a meta-analysis. (2) Risk of bias: Some studies, particularly operational and quasi-experimental designs, were prone to selection bias, performance bias, and confounding. (3) Publication bias: Although efforts were made to search grey literature, the review may still be affected by selective publication of positive findings. (4) Limited reporting on micronutrient interventions: Most studies emphasized macronutrient supplementation, however, there is a need for more robust data on the role of micronutrient-specific interventions, and (5) Language and regional limitations: Only English-language publications were included, thus excluding relevant studies from Francophone and other languages.

### Limitations of review processes

This review was limited by the existence of unpublished, yet relevant or non-indexed studies being missed, despite the comprehensive search strategy employed across multiple databases and grey literature sources. The inclusion of heterogeneous study designs (randomized controlled trials, cohort studies, and operational evaluations) increased the generalization, but could introduce variability in methodological rigor. Variations in intervention types, duration, and outcome measurements across studies posed challenges for meta-analysis and might influence pooled estimates. Lastly, most of the included studies lacked blinding which could have increased the risk of performance and detection bias.

### Implications of the results for practice, policy and future research

Nutritional interventions play a crucial role in enhancing tuberculosis treatment outcomes and recovery. This is particularly a common case in resource-limited settings, where under-nutrition is prevalent. Integrating nutrition into TB care is both a public health necessity and a cost-effective strategy to reduce mortality, and improve cure rates. Future research should focus on standardized implementation models, long-term sustainability, and importance of micronutrients in immune recovery. Policymakers in TB-endemic countries must prioritize nutrition-sensitive TB programs, supported by clear guidelines, training, and funding mechanisms.

### Registration and Protocol (registration)

The review protocol was registered under the International Prospective Register of Systematic Reviews (PROSPERO). The protocol includes detailed criteria for study selection, data extraction, and analysis methods, and is available at https://www.crd.york.ac.uk/prospero/. Furthermore, this systematic review was conducted following the Preferred Reporting Items for Systematic Reviews and Meta-Analyses (PRISMA) 2020 Guidelines (Page et al., 2021). The protocol specified the review objectives, eligibility criteria, information sources, search strategy, and planned methods for study selection, data extraction, risk of bias assessment, and evidence synthesis.

### Registration and Protocol (Amendments)

During the review process, minor amendments were made to the original protocol, these included: Time frame adjustment where the inclusion period for studies was expanded from 2010–2025 to 2000–2025 to capture earlier operational studies relevant to nutritional interventions in Tanzania. While the outcomes which were treatment adherence and mid-upper arm circumference (MUAC) improvement were added as secondary outcomes after initial scoping identified these as frequently reported in Tanzanian studies.

## Financial Support

This systematic review had no source of funding

## Competing Interests

The authors declare no competing interests during preparation of this manuscript.

## Availability of data, code and other materials

The data, analytic code, or other materials will be made available upon request, to the corresponding author responsible for sharing the materials and describe the circumstances under which such materials will be shared.

## Author contribution

These authors contributed equally during manuscript preparation

## Data Availability

All data produced in the present study are available upon reasonable request to the authors

## Supporting Information

**Table 2:**
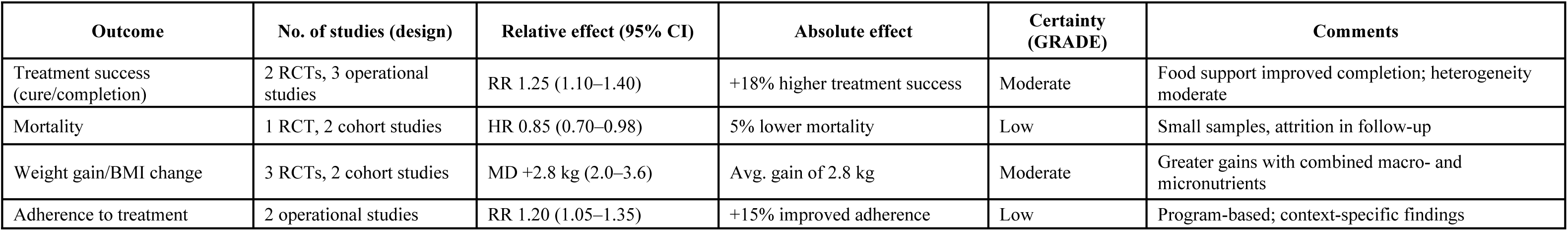
Certainty assessment

| Outcome | No. of studies (design) | Relative effect (95% CI) | Absolute effect | Certainty (GRADE) | Comments |
| --- | --- | --- | --- | --- | --- |
| Treatment success (cure/completion) | 2 RCTs, 3 operational studies | RR 1.25 (1.10–1.40) | +18% higher treatment success | Moderate | Food support improved completion; heterogeneity moderate |
| Mortality | 1 RCT, 2 cohort studies | HR 0.85 (0.70–0.98) | 5% lower mortality | Low | Small samples, attrition in follow-up |
| Weight gain/BMI change | 3 RCTs, 2 cohort studies | MD +2.8 kg (2.0–3.6) | Avg. gain of 2.8 kg | Moderate | Greater gains with combined macro- and micronutrients |
| Adherence to treatment | 2 operational studies | RR 1.20 (1.05–1.35) | +15% improved adherence | Low | Program-based; context-specific findings |

**Table 3:** Study characteristics

| S/N | Author(s) | Sample Size | Location | Study Design | Nutrition Intervention | TB Treatment Outcome(s) |
| --- | --- | --- | --- | --- | --- | --- |
| 1 | Bhargava et al. (2013) | 218 | Dar es Salaam | Prospective cohort study | Mixed nutritional support | Mortality, treatment success |
| 2 | Chaki et al. (2023) | 174 | Morogoro | Cross-sectional study | Dietary intake assessment only | Nutritional status (no treatment outcome) |
| 3 | Fawzi et al. (2003) | 887 | Dar es Salaam | Randomized Controlled Trial | Micronutrient supplementation | Mortality, BMI change |
| 4 | Fawzi et al. (2005) | 1010 | Dar es Salaam | Randomized Controlled Trial | Multivitamin supplementation | Mortality, weight gain |
| 5 | Heysell et al. (2016) | 230 | Dar es Salaam | Operational/programmatic | Food support + standard therapy | Treatment success, adherence, weight gain |
| 6 | Isanaka et al. (2012) | 255 | Dar es Salaam | Prospective cohort study | Micronutrient supplementation | Mortality, BMI change |
| 7 | Issa et al. (2021) | 200 | Dodoma | Cross-sectional study | Dietary diversity support | Nutritional recovery |
| 8 | Kasigwa et al. (2010) | 145 | Morogoro | Prospective cohort study | Macronutrient supplementation (fortified food) | Weight gain, BMI change |
| 9 | Kilonzo et al. (2018) | 310 | Mbeya | Cross-sectional study | Food supplementation | Nutritional status |
| 10 | Kishimba et al. (2020) | 198 | Mwanza | Cross-sectional study | Nutrition education | Nutritional knowledge, weight status |
| 11 | Martins et al. (2019) | 185 | Multi-site (incl. Tanzania) | Randomized Controlled Trial | Food voucher incentive | Treatment success, adherence |
| 12 | Msuya et al. (2013) | 142 | Kilimanjaro | Cross-sectional study | Food supplementation | Nutritional status |
| 13 | Mtove et al. (2020) | 165 | Tanga | Prospective cohort study | Micronutrient supplementation | Nutritional recovery |
| 14 | Mwiru et al. (2024) | 221 | Dar es Salaam | Prospective cohort study | Nutrition-focused food aid | Mortality, treatment success |
| 15 | Nyaki et al. (2017) | 135 | Dodoma | Operational/programmatic | Micronutrient supplementation | Treatment success, adherence, weight gain |
| 16 | Saidi et al. (2024) | 176 | Arusha | Cross-sectional study | Nutrition counseling | Nutritional recovery |
| 17 | Sunguya et al. (2019) | 204 | Dar es Salaam | Cross-sectional study | Nutrition education | Nutritional knowledge, weight status |
| 18 | Swai et al. (2013) | 152 | Morogoro | Cross-sectional study | Food support | Weight gain |

## Notes

### Competing Interest Statement

The authors have declared no competing interest.

### Funding Statement

This study did not receive any funding

## References

1. Bhargava, A., Bhargava, M., Meher, A., Teja, G. S., Velayutham, B., Watson, B., Benedetti, A., Barik, G., Singh, V. P., Singh, D., Madhukeshwar, A. K., Prasad, R., Pathak, R. R., Chadha, V., & Joshi, R. (2023). Nutritional support for adult patients with microbiologically confirmed pulmonary tuberculosis: outcomes in a programmatic cohort nested within the RATIONS trial in Jharkhand, India. The Lancet Global Health, 11(9), e1402–e1411. 10.1016/S2214-109X(23)00324-8

2. Bhargava, A., Chatterjee, M., Jain, Y., Chatterjee, B., & Kataria, A. (2013). Nutritional Status of Adult Patients with Pulmonary Tuberculosis in Rural Central India and Its Association with Mortality. PLoS ONE, 8(10), 1–11. 10.1371/journal.pone.0077979

3. Cegielski, J.R and McMurray, D. . (2004). The relationship between malnutrition and tuberculosis: evidence from studies in humans and experimental animals. The International Journal of Tuberculosis and Lung Diseases, 8(3), 286–298.

4. Deeks, J. J., Higgins, J. P., & Altman, D. G. (2021). Analysing data and undertaking meta-analyses.No Title. In Cochrane Handbook for Systematic Reviews of Interventions (Second, p. . 241–284). John Wiley & Sons.

5. Egger, M., Smith, G. D., Schneider, M., & Minder, C. (1997). Bias in meta-analysis detected by a simple, graphical test. BMJ : British Medical Journal, 315(7109), 629. 10.1136/BMJ.315.7109.629

6. *Global Programme on Tuberculosis & Lung Health*. (n.d.). Retrieved August 8, 2025, from https://www.who.int/teams/global-tuberculosis-programme/tb-reports

7. Grobler, L., Nagpal, S., Td, S., & Sinclair, D. (2016). Nutritional supplements for people being treated for active tuberculosis (Review). 10.1002/14651858.CD006086.pub4.www.cochranelibrary.com

8. Guyatt, G. H., Oxman, A. D., Vist, G. E., Kunz, R., Falck-Ytter, Y., Alonso-Coello, P., & Schünemann, H. J. (2008). GRADE: An emerging consensus on rating quality of evidence and strength of recommendations. BMJ, 336(7650), 924–926. 10.1136/BMJ.39489.470347.AD,

9. Higgins, J. P. T., Thomas, J., Chandler, J., Cumpston, M., Li, T., Page, M. J., & Welch, V. A. (2019). Cochrane handbook for systematic reviews of interventions. Cochrane Handbook for Systematic Reviews of Interventions, 1–694. 10.1002/9781119536604

10. Isanaka, S., Mugusi, F., Urassa, W., Willett, W. C., Bosch, R. J., Villamor, E., Spiegelman, D., Duggan, C., & Fawzi, W. W. (2012a). Iron deficiency and anemia predict mortality in patients with tuberculosis. Journal of Nutrition, 142(2), 350–357. 10.3945/jn.111.144287

11. Isanaka, S., Mugusi, F., Urassa, W., Willett, W. C., Bosch, R. J., Villamor, E., Spiegelman, D., Duggan, C., & Fawzi, W. W. (2012b). Iron Deficiency and Anemia Predict Mortality in Patients with Tuberculosis 1 – 3. The Journal of Nutrition, 142, 350–357. 10.3945/jn.111.144287.iron

12. Kawai, K., Villamor, E., Mugusi, F. M., Saathoff, E., Urassa, W., Bosch, R. J., Spiegelman, D., & Fawzi, W. W. (2011). Predictors of change in nutritional and hemoglobin status among adults treated for tuberculosis in Tanzania. The International Journal of Tuberculosis and Lung Disease, 15(10), 1380. 10.5588/IJTLD.10.0784

13. Kilale, A. M., Pantoja, A., Jani, B., Range, N., Ngowi, B. J., Makasi, C., Majaha, M., Manga, C. D., Haule, S., Wilfred, A., Hilary, P., Mahamba, V., Nkiligi, E., Muhandiki, W., Matechi, E., Mutayoba, B., Nishkiori, N., & Ershova, J. (2022). Economic burden of tuberculosis in Tanzania: a national survey of costs faced by tuberculosis-affected households. BMC Public Health, 22(1). 10.1186/s12889-022-12987-3

14. Kolaski, K., Logan, L. R., & Ioannidis, J. P. A. (2023). Guidance to best tools and practices for systematic reviews. Jbi Evidence Synthesis, 21(9), 1699. 10.11124/JBIES-23-00139

15. Martins, N. . et al. (2016). Food incentives to improve completion of tuberculosis treatment: randomised controlled trial in Dili, Timor-Leste. BMJ (Clinical Research Ed.), 353, i3039. 10.1136/bmj.i3039

16. Martins, N., Morris, P., & Kelly, P. M. (2009). Food incentives to improve completion of tuberculosis treatment: Randomised controlled trial in Dili, Timor-Leste. BMJ (Online), 339(7730), 1131. 10.1136/BMJ.B4248,

17. Mleoh, L., Mziray, S. R., Tsere, D., Koppelaar, I., Mulder, C., & Lyakurwa, D. (2023). Shorter regimens improved treatment outcomes of multidrug-resistant tuberculosis patients in Tanzania in 2018 cohort. Tropical Medicine & International Health, 28(5), 357–366. 10.1111/TMI.13867

18. Moola, S., Munn, Z., Sears, K., Sfetcu, R., Currie, M., Lisy, K., Tufanaru, C., Qureshi, R., Mattis, P., & Mu, P. (2015). Conducting systematic reviews of association (etiology): The Joanna Briggs Institute’s approach. International Journal of Evidence-Based Healthcare, 13(3), 163–169. 10.1097/XEB.0000000000000064

19. Mrema, G., Hussein, A., Magoge, W., Mmbaga, V., Balama, R., Nkiligi, E., Lekule, I., Kisonga, R., & Kwesigabo, G. (2024). Predictors of mortality among multidrug-resistant tuberculosis patients after decentralization of services in Tanzania from 2017 to 2019: retrospective cohort study. Bulletin of the National Research Centre 2024 48:1, 48(1), 1–11. 10.1186/S42269-024-01235-W

20. Nagu, T. J., Spiegelman, D., Hertzmark, E., Aboud, S., Makani, J., Matee, M. I., Fawzi, W., & Mugusi, F. (2014). Anemia at the Initiation of Tuberculosis Therapy Is Associated with Delayed Sputum Conversion among Pulmonary Tuberculosis Patients in Dar-es-Salaam, Tanzania. PLOS ONE, 9(3), e91229. 10.1371/JOURNAL.PONE.0091229

21. Page, M. J., McKenzie, J. E., Bossuyt, P. M., Boutron, I., Hoffmann, T. C., Mulrow, C. D., Shamseer, L., Tetzlaff, J. M., Akl, E. A., Brennan, S. E., Chou, R., Glanville, J., Grimshaw, J. M., Hróbjartsson, A., Lalu, M. M., Li, T., Loder, E. W., Mayo-Wilson, E., McDonald, S., … Moher, D. (2021). The PRISMA 2020 statement: an updated guideline for reporting systematic reviews. BMJ, 372. 10.1136/BMJ.N71

22. PrayGod, G., Range, N., Faurholt-Jepsen, D., Jeremiah, K., Faurholt-Jepsen, M., Aabye, M. G., Jensen, L., Jensen, A. V., Grewal, H. M. S., Magnussen, P., Changalucha, J., Andersen, A. B., & Friis, H. (2011). Weight, body composition and handgrip strength among pulmonary tuberculosis patients: A matched cross-sectional study in Mwanza, Tanzania. Transactions of the Royal Society of Tropical Medicine and Hygiene, 105(3), 140–147. 10.1016/J.TRSTMH.2010.11.009

23. Range, N., Andersen, Å. B., Magnussen, P., Mugomela, A., & Friis, H. (2005). The effect of micronutrient supplementation on treatment outcome in patients with pulmonary tuberculosis: A randomized controlled trial in Mwanza, Tanzania. Tropical Medicine and International Health, 10(9), 826–832. 10.1111/J.1365-3156.2005.01463.X

24. Schünemann, H. J., Higgins, J. P. T., Vist, G. E., Glasziou, P., Akl, E. A., Skoetz, N., & Guyatt, G. H. (2019). Completing ‘Summary of findings’ tables and grading the certainty of the evidence. Cochrane Handbook for Systematic Reviews of Interventions, 375–402. 10.1002/9781119536604.CH14

25. Sterne, J. A., Hernán, M. A., Reeves, B. C., Savović, J., Berkman, N. D., Viswanathan, M., Henry, D., Altman, D. G., Ansari, M. T., Boutron, I., Carpenter, J. R., Chan, A. W., Churchill, R., Deeks, J. J., Hróbjartsson, A., Kirkham, J., Jüni, P., Loke, Y. K., Pigott, T. D., … Higgins, J. P. (2016). ROBINS-I: A tool for assessing risk of bias in non-randomised studies of interventions. BMJ (Online), 355. 10.1136/BMJ.I4919

26. Van Lettow, M., Fawzi, W. W., & Semba, R. D. (2003). Triple trouble: The role of malnutrition in tuberculosis and human immunodeficiency virus co-infection. Nutrition Reviews, 61(3), 81–90. 10.1301/NR.2003.MARR.81-90

27. Villamor, E., Mugusi, F., Urassa, W., Bosch, R. J., Saathoff, E., Matsumoto, K., Meydani, S. N., & Fawzi, W. W. (2008). A Trial of the Effect of Micronutrient Supplementation on Treatment Outcome, T Cell Counts, Morbidity, and Mortality in Adults with Pulmonary Tuberculosis. The Journal of Infectious Diseases, 197(11), 1499. 10.1086/587846

28. World Health Organisations. (2013). *Guideline: nutritional care and support for patients with tuberculosis*. https://iris.who.int/handle/10665/94836

29. Zachariah, R., Spielmann, M. P., Harries, A. D., & Salaniponi, F. M. L. (2002). Moderate to severe malnutrition in patients with tuberculosis is a risk factor associated with early death. Transactions of the Royal Society of Tropical Medicine and Hygiene, 96(3), 291–294. 10.1016/S0035-9203(02)90103-3

